# Longitudinal study about low back pain, mental health, and access to healthcare system during COVID-19 pandemic: protocol of an ambispective cohort

**DOI:** 10.1101/2020.07.22.20160309

**Authors:** Natan Feter, Eduardo L Caputo, Igor R Doring, Jayne S Leite, Júlia Cassuriaga, Felipe F Reichert, Marcelo C da Silva, Airton J Rombaldi

## Abstract

This study aims to investigate the effects of physical activity before, during, and after social distancing due to the COVID-19 pandemic on low back pain (LBP), mental health and healthcare access. The PAMPA Cohort (**P**rospective Study **A**bout **M**ental and **P**hysical He**a**lth) is a state-level ambispective longitudinal observational study that will be conducted in Rio Grande do Sul, Brazil. An online-based questionnaire will be used to assess LBP, mental health, healthcare access and physical activity at four time points: 1) pre-COVID-19 social distancing, 2) during COVID-19 social distancing, 3) 6 months and 4) 12 months after baseline. A proportional sample size calculation was conducted, and the final sample size was estimated in 1,767 people, distributed in seven state regions. Participants will be recruited by a four-arm approach: contact with universities, social media, local media and personal contacts. Descriptive analyzes will be reported as mean or proportion and respective 95% confidence interval (CI), when appropriate. Comparison between pre- and during COVID-19 social distancing, and after baseline assessments will be performed using two-way ANOVA with repeated measures. Proportions will be compared by Chi-squared test.

## Introduction

In December of 2019, the first cases of the new coronavirus disease (COVID-19) were documented. Three months later, the World Health Organization (WHO) declared the COVID-19 a pandemic[1] with almost 9 million cases worldwide in mild June of 2020 [2]. The rapid spreading velocity of this virus[3] led robust healthcare systems, such as in Germany and Great Britain to a chaotic scenario[4]. Although an unprecedented effort has been made to develop an effective treatment and cure for COVID-19, nothing was discovered to date. In this scenario, public health strategies to prevent virus spreading needed to be implemented. The non-pharmacological strategies that have been proved as the most effective mechanism to reduce virus spreading are social distancing, use of mask, and quarantine. Social distancing aims to reduce interaction between individuals to reduce both velocity and rate of infection and then protecting the healthcare system against a collapse[5]. This strategy is carried out by interrupting all non-essential activities and services, such as schools and gyms, and canceling sports events and concerts [6].

Although social distancing is the best strategy to reduce new cases of the COVID-19, it might be followed by health-related side effects such as musculoskeletal pain, physical inactivity, poor mental health, and economic challenges at both individual and community-level[7]. Low back pain (LBP), affects 7.3% of world population and is one of the main causes of physical disability and days of work lost [8, 9]. Social distancing might lead people to longer sitting time[10] which is associated with LBP [11, 12]. Also, physical activity has shown beneficial effects on LBP and other disabilities, which highlight the importance of being physically active[13]. These associations enforce the importance of promoting physical activity during social distancing to reduce time spent on sedentary activity and LBP [14, 15].

Similarly, mental health was severally impacted at global level during the COVID-19 pandemic. Misleading information about virus transmission, vaccines and treatments development, and cumulative cases of people who have COVID-19 have led to anxiety, fear, and hopelessness[16]. Besides, increased time spent in sedentary activities[17, 18] and decreased participation in moderate-to-vigorous physical activities[19, 20] may be associated with higher incidence of depression and anxiety. In order to mitigate the impact of the COVID-19 crisis in global mental health, WHO launched in March 2020 a set of considerations to protect mental health for general population, healthcare workers, careers, older adults, and isolated people. Further, physical activity is a known non-pharmacologic, low-cost strategy to reduce the risk for depression[21] and anxiety[22]. Failure in acting seriously and promptly to protect mental health of whole-population during COVID-19 pandemic will lead to an even greater economic, social, and health crisis[23].

Another impact of social distancing strategy in population-level health outcomes is on healthcare access. A multinational survey conducted by WHO in 159 countries revealed that in 72% of high-income countries, the COVID-19 preparedness plan included strategies to provide health services for non-communicable chronic diseases (NCD) patients, while this proportion was 42% in low-income countries[24]. The same survey showed that half of the patients with hypertension or diabetes/diabetes-related complications had their treatments partially or completely disrupted during the COVID-19 pandemic. To mitigate this lack of access, telemedicine has been adopted in 58% of countries worldwide to support people with NCD to continue receiving treatment. However, in low- and middle-income countries, where 85% of all death by NCD occur, only 4 out of 10 countries have offered this service. Then, it is vital to ensure that people with NCD have access to treatment and prescribed medicines to reduce the impact of the COVID-19 in these patients especially in low-and middle-income countries.

Based on the impact of COVID-19 on different aspects of physical and mental health, the present study aims to (1) investigate the effect of physical activity before, during, and after social distancing due to COVID-19 pandemic in LBP and mental health and (2) identify the impact of social distancing in healthcare access.

## Methods

### Study design

The study protocol was approved by the institutional research ethics board of the Superior School of Physical Education of the Federal University of Pelotas, Brazil (protocol: 4.093.170). The PAMPA cohort (**P**rospective Study **A**bout **M**ental and **P**hysical He**a**lth) will be characterized as an ambispective longitudinal observational study. A questionnaire will be used to gather data on mental and physical health, as well as healthcare access at four time points: 1) pre-COVID-19 social distancing, 2) during COVID-19 social distancing, 3) 6 months and 4) 12 months after baseline assessments, as shown in Figure 1.

**Figure 1.**
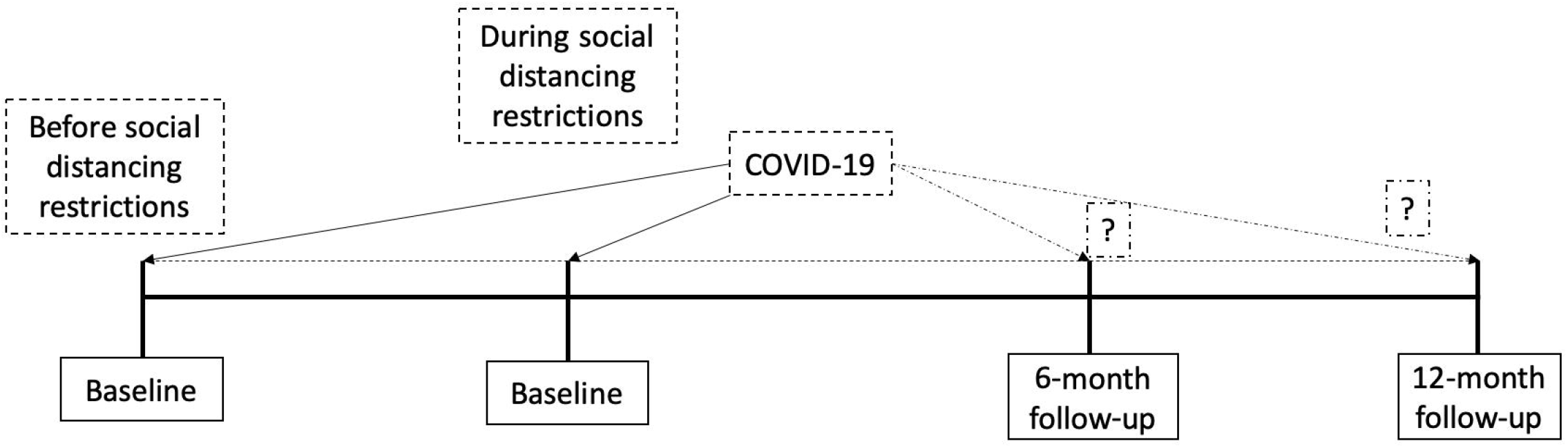
The PAMPA Cohort follow-up schedule.

### Sample

Sample size was calculated based on our three primary outcomes prevalence (i.e. LBP, mental health, and healthcare access). According to the latest Brazilian National Health Survey[25], Rio Grande do Sul state has the highest prevalence of depression amongst all states (13.2%; 95%CI: 11.8%-15.0%). Also, the same survey reported that 86.0% (95%CI: 83.7%-88.1%) of people in the state was able to access drugs prescribed in the last health care visit, two weeks prior the survey[26]. Finally, in 2011, Ferreira et al.[27] reported a LBP prevalence of 40% (95%CI: 36.9% to 43.2%) in a southern Brazil city. The 2010 populational census in Brazil reported a total population size of 10,693,929 people living in Rio Grande do Sul[28]. The prevalence of depression required the highest sample size (depression: N=1,359; LBP: N=960; healthcare and medication access: N=820) to ensure a 95% confidence interval and 1.8 margin of error. Also, we proportionally distributed this sample size along six different mesoregions within the state. Further, we accounted for a possible lost-to-follow-up up to 30%. Therefore, our final sample size was estimated in 1,767 and Figure 2 indicates the target number of participants by the state mesoregions.

**Figure 2.**
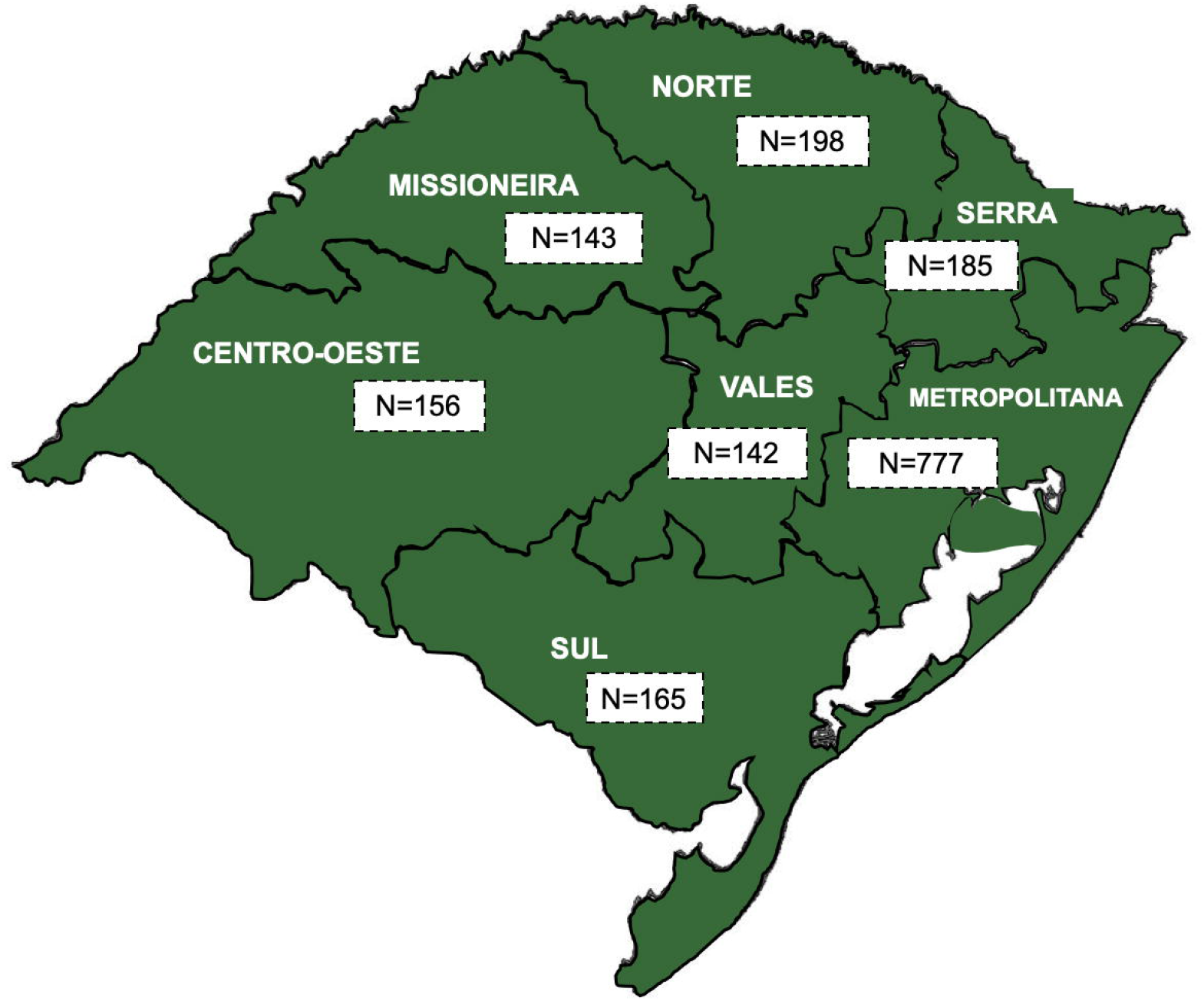
Target sample size according to different mesoregions.

### Questionnaire

We developed an online-based, self-administered questionnaire to evaluate the effects of physical activity before, during, and after social distancing on mental health, LBP, and other health outcomes. The questionnaire was developed using the Google® Forms platform. A preliminary analysis showed that the average time to complete the survey was around 10 minutes, ranging from 7 to 12 minutes. Questions about mental health, LBP, and physical activity will be asked twice to access these outcomes based on different time periods (before and during social distancing). In the first moment, questions are based on pre-social distancing while the second are based on the current moment (i.e. during social distancing).

### Informed consent form

The first question is whether the participant accepts to participate in the study. If participant’s answer is “No”, the form will be automatically uploaded without any participant’s information. If participant’s answer is “Yes”, the definitive questionnaire will be prompted to participant’s screen in the next page.

### Socioeconomic information

Questions on age, gender (male, female, other/prefer not to mention), city of residence, ethnicity (white, black, yellow, mixed, indigenous, other), marital status (with or with no partner) and number of people living in the same household will be asked. The highest educational level achieved and current job will also be assessed.

### Economic impact of COVID-19 pandemic

The impact of COVID-19 pandemic in work-related activities and economic situation will be evaluated. Questions regarding home-office adaptation, working hours before and during pandemic, and whether social distancing affected participant’s income will be addressed.

### COVID-19: knowledge and attitudes

Self-rated knowledge about the COVID-19 will be rated as “bad”, “regular”, “good”, “very good”, or “excellent”. Attitudes towards social distancing will be asked by two questions. The first is “*Regarding the social distancing that is being guided by health authorities, that is, staying home and avoiding contact with other people, how much of it do you think you are managing to do?*”. The options available are “very little”, “little”, “somewhat”, “very much” and “totally isolated”. The second question is “*How has your daily routine at home been?*”. Answering options are: “staying at home all the time”, “go out only for essential things” (e.g. buying food, go out sometimes to shopping and stretching my legs), “go out every day for some activity”, or “go out every day to work or another regular activity”. We further ask two questions about participants opinion on social distancing and flexibilization of it (e.g. re-opening of schools, gyms, markets).

### Primary outcomes

#### Mental health

Mental health was assessed by the Hospital Anxiety and Depression Scale (HDAS), which identified symptoms of depression and anxiety in both pre- and during social distancing. This 14-item scale was designed to provide a simple and reliable tool to be used in both community settings and primary care medical practice[29]. Each domain (depression and anxiety) has seven items that are scored between 0 and 3. Therefore, each domain has a maximum score of 21. Participants who scored less than 7 will be classified as non-cases for that domain. Scores between 8 and 10 will be considered as mild cases, between 11 and 14 as moderate, and between 15 and 21 as severe cases of depression and/or anxiety[30]. Further, we added a single question about self-rated memory to provide an indication of whether the respondent is worried about his/her memory. They will be asked to rate whether their memory at the present time as “excellent”, “very good”, “good”, “fair” or “poor”.

#### LBP

LBP experience, as well as activity limitation, pain intensity and care seeking will be assessed. An image of a person in the supine position with the low back area highlighted in a different color will be used together with the LBP experience question, as follow: “*Before social isolation have you had pain in your lower back, as shown in the figure, for at least one day?”*. Pain intensity will be assessed using a numeric pain rating scale as “0” indicating no pain and “10” indicating the worst pain possible. Activity limitation related to LBP will be assessed by asking: “*Before social isolation was your low back pain strong enough to limit or alter your daily activities for at least one day?”*. Participants will be also asked if they seek care for the following healthcare professionals to manage their LBP: “General practitioner”, “Physiotherapist”, “Exercise Science Professional” and “Other”. Questions related to the second time point (during COVID-19 social distancing) has the same format; however, the time-reference will be the current week.

#### Healthcare access

The participants will be asked about the diagnostic of chronic diseases based on a question used in the Brazilian Surveillance System of Risk Factors for Chronic Diseases by Telephone Interviews (VIGITEL)[31]. If the answer was “Yes”, the participants will be asked about medicine and medical assistance needed and access to these services. Also, we will investigated about the type of health service (public or private), and the reason to seek or to withdraw with it. Furthermore, we will measure the perception about social isolation on chronic disease control (much worse, worsened, not changed, has improved, has improved a lot).

### Exposure

#### Physical activity

Physical activity before and during the social distancing period will be assessed by two similar questions. The first one is about physical activity practiced before the COVID-19 social distancing: “*Before social distancing, were you engaged in physical activity regularly?*”[32]. If participant’s answer is “Yes”, the total days and duration (minutes) that he/she performed physical activity in a regular pre-COVID-19 week will be asked. The second question has the same format; however, the time-reference will be the current week (during COVID-19 pandemic social distancing). In addition, we will ask what type of physical activity participants usually engage.

#### Participant recruitment

To achieve the target sample size (N=1,767), we will use a four-arm approach, as illustrated below (Figure 3). The progress of data collection in each mesoregion will be followed on a daily basis. Further, researches will meet every Monday, Wednesday and Friday to follow daily progress and adjust, if necessary, recruitment strategies.

**Figure 3.**
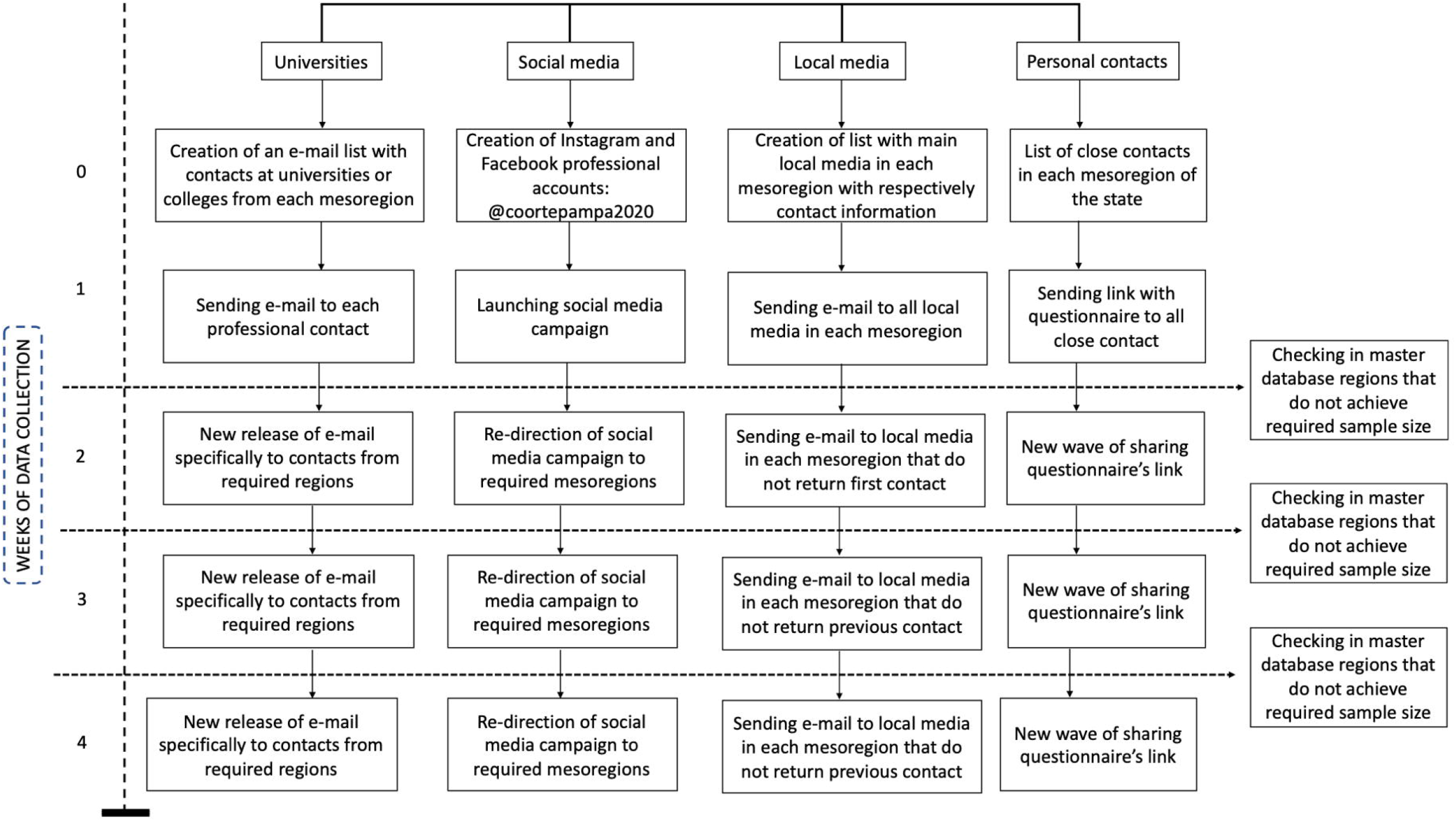
Schematic recruitment strategy of the PAMPA Cohort.

In the week before data collection begins, each researcher provided a list of possible contacts from universities or colleges in each mesoregion. A standardized message was sent to these contacts by email, with information about the survey objectives, identification of the researcher’s coordinators, and a link to access the questionnaire. Social media campaigns at Instagram® and Facebook® will be used to spread the questionnaire’s link to different regions within the state. Besides that, daily publications on research’s pages in both social medias will be planned to reach more people. Every week, the campaign will be adjusted as necessary. Also, local media will be contacted by email and social media to inform local population about the present study. Finally, each researcher involved in this survey will share the link with the questionnaire access to personal contacts spread across the state. Recruitment phase will have a total duration of four weeks.

## Data analyzes

Data will be exported from Google® Sheets to a Microsoft® Excel spreadsheet. Then, data will be exported to Stata 13.1, where analyzes will be carried out. Descriptive analyzes will be performed, and data reported as mean or proportion and respectively 95% confidence interval (CI), when appropriate. Proportions will be compared by Chi-squared test. Comparison between pre-, during, and after COVID-19 social distancing will be performed using two-way ANOVA with repeated measures. Groups will be defined according to physical activity level based on the World Health Organization recommendation[33]: physically inactive (less than 150 minutes of moderate-to-vigorous physical activity [MVPA] per week) and active (150 minutes or more of MVPA per week). Time will be referenced as the period of data collection (pre, during, and after social isolation). Group × time interaction will be also tested. Bonferroni’s post hoc will be used as required.

## Data Availability

Data wil be made available upon reasonable request.

